# Are medical students aware of SUDEP? A survey in Turkey

**DOI:** 10.1101/2023.07.14.23292665

**Authors:** Feyza Sule Aslan, Aylin Ismayilova, Shams Hasanli, Efthalia Angelopoulou, Kursad Nuri Baydili, Enes Akyuz

**Affiliations:** International School of Medicine, University of Health Sciences, Istanbul, Turkey; Department of Biological Chemistry, Medical School, National and Kapodistrian University of Athens, Athens, Greece; Department of Biostatistics, School of Medicine, University of Health Sciences, Istanbul, Turkey; Department of Biophysics, International School of Medicine, University of Health Sciences, Istanbul, Turkey

**Keywords:** sudden unexpected death in epilepsy, seizure, medical education

## Abstract

**Objective:** Sudden unexpected death in epilepsy (SUDEP) has been recognized as an important cause of death in patients with epilepsy. In order to inform patients with epilepsy and their relatives correctly, it is necessary to increase the awareness of students about SUDEP from the early stages of medical education. The aim of this study was to identify the level of knowledge and awareness of medical students in Turkey about SUDEP.

**Methods:** Medical students (23±7 years old; n=793) in Turkey participated in the online SUDEP awareness survey. The survey included demographic evidence, followed by questions about their awareness of epilepsy, seizure knowledge and about the definition, awareness of SUDEP.

**Results:** The majority of medical students (95%) claimed that they had heard about epileptic seizures. Half of the participants (49.9%) mentioned that they had heard about tonic-clonic seizures. However, two-thirds of the students (67%) have never heard about SUDEP, while 85% of the students stated that they did not have sufficient knowledge about SUDEP. Concerning the potential prevention of SUDEP, 80.8% of the students did not know about this topic. Furthermore, most participants (82%) expressed their interest and willingness to learn about SUDEP.

**Conclusion:** Knowledge about SUDEP plays a key role in identifying patients at risk and informing patients and their relatives. The limited awareness of SUDEP in medical education may pose risks for patients diagnosed with epilepsy and their relatives, and the effective incorporation of lectures and training in SUDEP into the curriculum of medical school is of paramount importance.

## 1. Introduction

Sudden Unexpected Death in Epilepsy (SUDEP) is defined as an unexpected and non-accidental death in patients diagnosed with epilepsy without neuropathological complications determined at the autopsy [1]. Although generalized tonic-clonic seizures (GTCS) and respiratory dysfunction are known risk factors, the pathophysiological mechanisms of SUDEP still remain unclear [2]. In addition, despite the increasing number of studies in SUDEP risk factors and causes, the research on the determination of the current level of knowledge and awareness of SUDEP among physicians and medical students is insufficient [3].

Research on how patients with epilepsy and their relatives should be informed about rare conditions such as SUDEP is rather controversial and relative studies use different approaches. Routine counseling regarding SUDEP in clinical practice still reflects an ongoing debate in the neurology community. Published guidelines and statements from professional organizations suggest routine counseling on SUDEP [4]. However, literature evidence shows that only a small minority of neurologists routinely counsel and inform their patients about the risk of SUDEP [5]. One of the main reasons for this choice of clinicians is the fear of creating anxiety and stress in patients after discussions about SUDEP [6].

Effective prevention of SUDEP requires a good knowledge of SUDEP and the risk factors. Therefore, the National Institute for Health and Care Excellence (NICE), the Scottish Intercollegiate Guidelines Network, and the American Epilepsy Association advocate discussing SUDEP with clinicians, patients, and families early in the clinical course of the medical condition to take preventive measures [7]. Brodie et al. recommended an individualized approach to discuss SUDEP as patients are ready to receive the information [8]. The reason for this recommendation is that patients at low risk of SUDEP (e.g., well-controlled seizures) or low-risk syndromes (e.g., childhood epilepsy) may face unnecessary distress. Therefore, physicians should be aware of the clinical entity of SUDEP, its definition, the risk factors and current preventive measures.

Although patients diagnosed with epilepsy initially fear SUDEP, most agree that the awareness of SUDEP motivates them to better manage the factors that trigger seizures. Awareness of SUDEP has increased among neurologists, researchers, patients diagnosed with epilepsy, and relatives of the patients. However, there are still uncertainties about if and to what extent non-physicians and especially medical students are aware of SUDEP [9]. In this study, we aimed to determine SUDEP knowledge of medical students in different periods of the 6 years of medical school via the SUDEP-awareness questionnaire.

## 2. MATERIAL and METHOD

### 2.1. Participants

Medical students aged 23±7 (n=793) studying at the medical school were asked to anonymously participate in an online survey after their informed consent. Medical students from different universities (67 university including University of Health Sciences, Tekirdağ Namık Kemal University, İstanbul University, Bursa Uludağ University etc.) in Turkey took part in the study. This study was approved by the Health Sciences University Hamidiye Scientific Research Ethics Committee with registration number 21/737.

The survey ensured that the students answered the questions by their own free will. Those who chose the “no” option at the beginning of the survey was excluded (refused to participate in the study). Questionnaires filled by students studying in other than medical faculties were also excluded.

### 2.2. Survey

The questionnaire used in the study was taken from the article named “Knowledge of sudden unexpected death in epilepsy (SUDEP) among 372 patients attending a German tertiary epilepsy center” by Surges et al. The survey was carried out in 2021 November-2022 April and lasted 6 months. The survey, which was organized via Google Form, was delivered to students online including e-mail, text message, medical student groups on WhatsApp, Instagram, Facebook. The survey had two parts: demographic and SUDEP-related questions. Demographic questions included age (age groups: 16-20, 21-25 or 26-30), gender, university they are studying, and year of medical school (year:1-6). In addition, students were asked whether they were interested in neurology (possible answers: yes, or no), whether they knew anyone with epilepsy, and if any family member had epilepsy. Also, students were also asked if there is anyone in their family who has heart or respiratory problems. In regard to their general knowledge about epilepsy, questions included “I know epilepsy”, “I know what a seizure is” and “I know what a tonic-clonic seizure is” (possible answers: yes, or no).

In the SUDEP questionnaire, medical students were asked whether they had heard of SUDEP before (possible answers: yes, or no), via which sources they had learned about SUDEP (possible answers: doctor, internet, written and visual media, or other sources), and their willingness to learn (more) about SUDEP (possible answers: yes, no or I don’t know). Finally, medical students were asked about quality of life in epilepsy and possible SUDEP treatment. (possible answers: yes, no, or I do not know).

### 2.3. Statistical analysis

Data analysis was carried out using the SPSS 25 package program. Frequencies and percentage values are presented for qualitative variables. Chi-square test was used for comparisons between independent qualitative variables. Type I error rate was accepted as 0.05 in the study.

## 3. RESULTS

Responses of the students (n=809) were obtained shortly after the survey and participants not meeting the inclusion criteria were excluded in this study (n:16). From the medical students included in the study (n=793), 33% were male (n=262) and 67% were female (n=531). Of these participants, 347 students were between the ages of 16-20; 429 were between the ages of 21-25 and 17 were between the ages of 26-30 (Table 1). According to the results of the survey, 95% (n=755) of the students had seizure awareness while 49.9% (n=398) had tonic-clonic seizure awareness. Only 47.8% of the students had knowledge about epilepsy (Figure 1, Table 2).

**Fig. 1.**
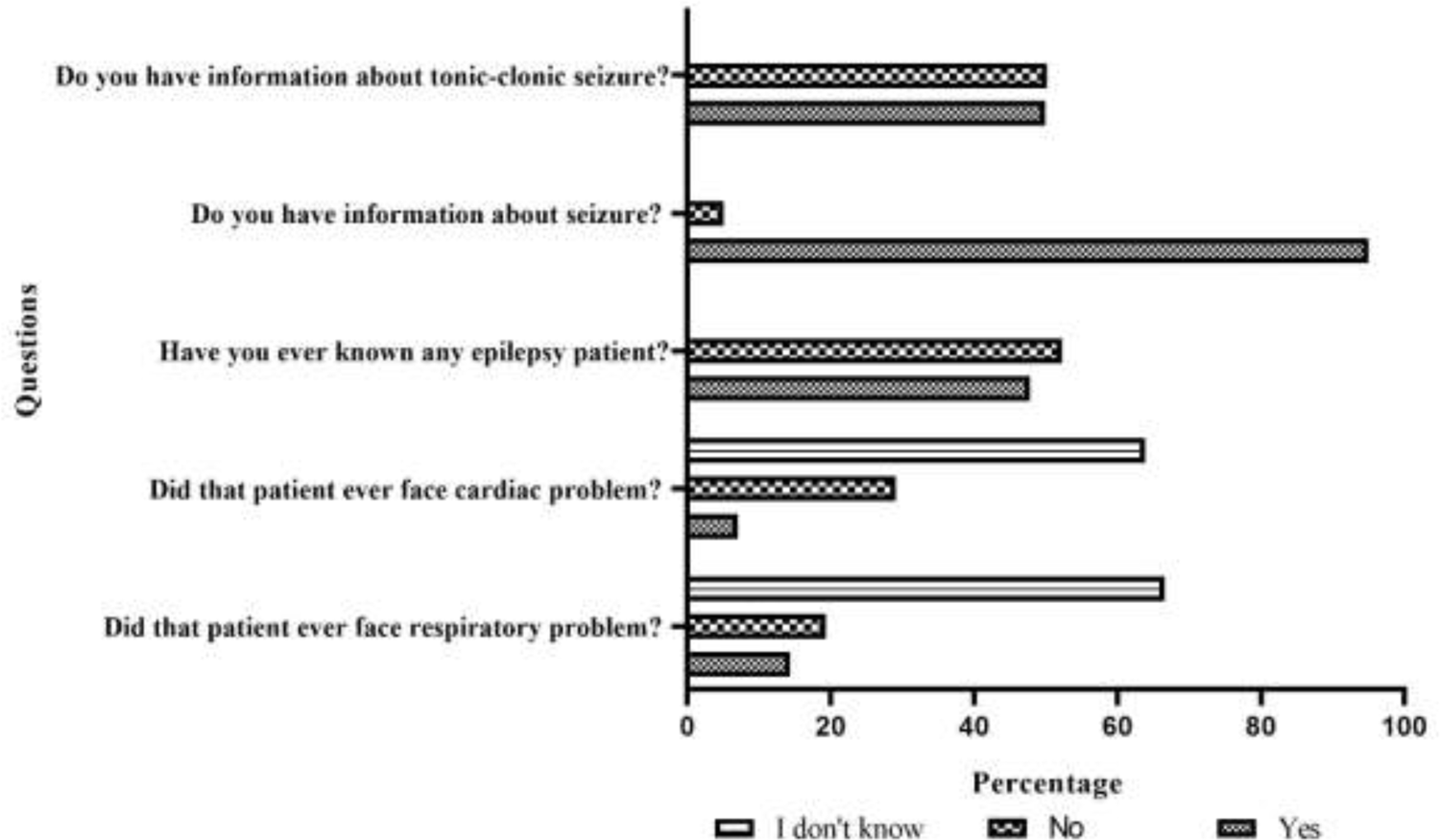
Answers of the participants for epilepsy-related questions. Most medical students had known an epilepsy patient and had been informed about seizures and tonic-clonic seizures.

**Table 1.** Demographic characteristics.

| Characteristics | Percentage (%) | Number (n) |
| --- | --- | --- |
| **Year of medical school** |  |  |
| 1 | 20,7 | 164 |
| 2 | 21,8 | 173 |
| 3 | 18,4 | 146 |
| 4 | 16,4 | 130 |
| 5 | 11,6 | 92 |
| 6 | 11,1 | 88 |
| **Age (years)** |  |  |
| 16-20 | 43,8 | 347 |
| 21-25 | 54,1 | 429 |
| 26-30 | 2,1 | 17 |
| **Sex** |  |  |
| Male | 33 | 262 |
| Female | 67 | 531 |
| **Marriage status** |  |  |
| Single | 98,7 | 783 |
| Married | 1,3 | 10 |

**Table 2.**
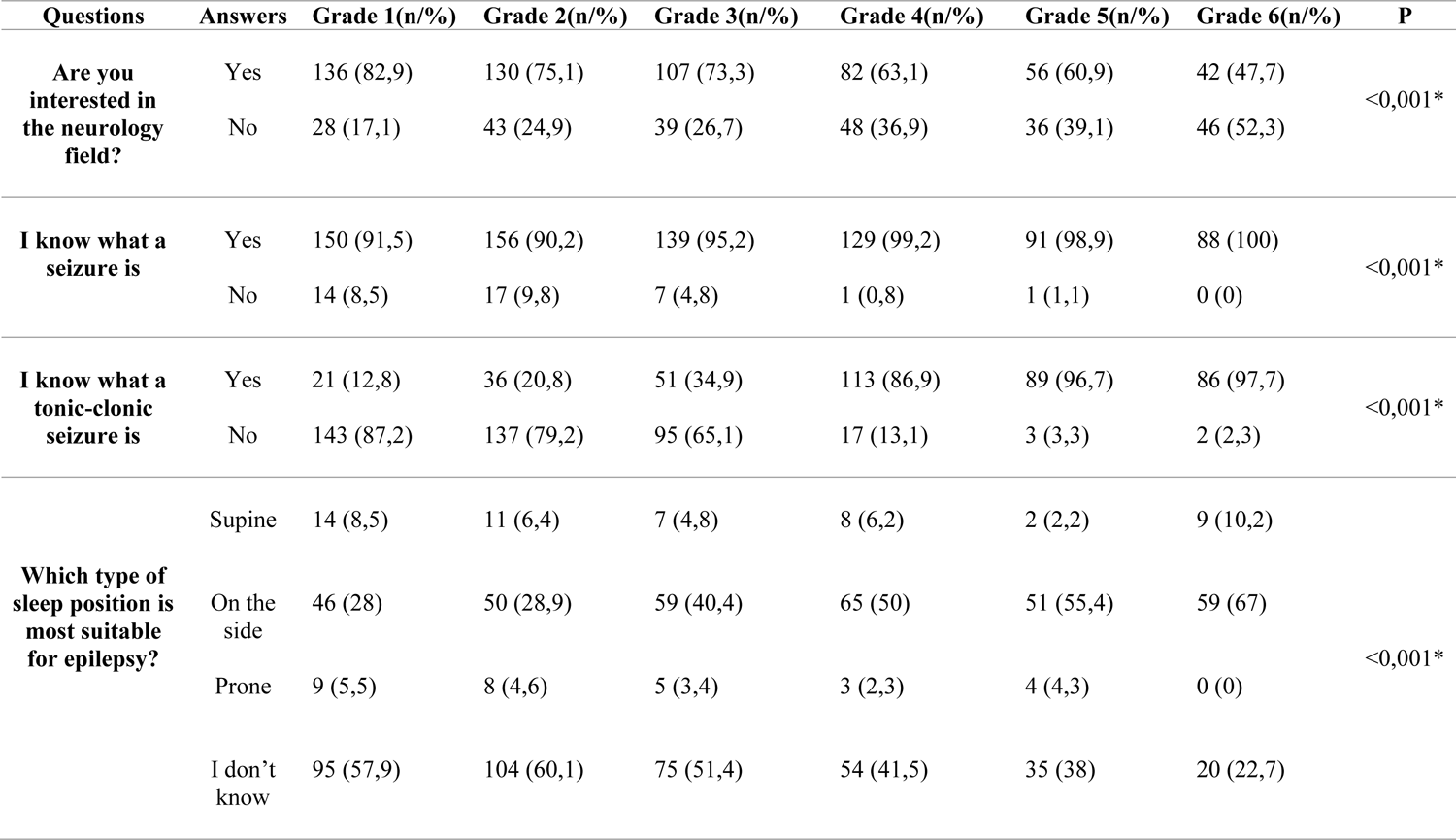
Students’ information about the SUDEP questionnaire according to the term.

According to the results, 85.5% of the students answered “No” to the question asked, *“Is there a history of epilepsy in your family?”* while the answer of “Yes” was 10.7%. The number of students who knew patients with epilepsy was 379 (47.8%). These students who knew the patients answered the questions *“Does this patient have cardiac and respiratory problems?”*.

Based on the results, the number of patients with epilepsy accompanying cardiac problems was reported as 44 (7.1%), and the number of patients with epilepsy having respiratory problems as 88 (14.3%). As a result, respiratory and cardiac problems were accompanied by patients with epilepsy recognized by the students. In addition, the classification of these answers were classified in Table 3.

**Table 3.** Evaluate the presence of people with epilepsy around the students from SUDEP questionnaire according to the term.

| Questions | Answers | Grade 1(n/%) | Grade 2(n/%) | Grade 3(n/%) | Grade 4(n/%) | Grade 5(n/%) | Grade 6(n/%) | P |
| --- | --- | --- | --- | --- | --- | --- | --- | --- |
| **Have you ever known any epilepsy patient?** | Yes | 75 (45,7) | 76 (43,9) | 63 (43,2) | 61 (46,9) | 52 (56,5) | 52 (59,1) | <0,076 |
|  | No | 89 (54,3) | 97 (56,1) | 83 (56,8) | 69 (53,1) | 40 (43,5) | 36 (40,9) |  |
| **Did that patient ever face cardiac problem?** | Yes | 7 (5,3) | 14 (10,3) | 6 (5,6) | 4 (4,2) | 6 (8,3) | 7 (9,5) | <0,214 |
|  | No | 34 (25,6) | 35 (25,7) | 28 (26,2) | 31 (32,3) | 22 (30,6) | 30 (40,5) |  |
|  | I don't know | 92 (69,2) | 87 (64) | 73 (68,2) | 61 (63,5) | 44 (61,1) | 37 (50) |  |
| **Did that patient ever face respiratory problem?** | Yes | 23 (17,3) | 25 (18,4) | 16 (15) | 9 (9,5) | 5 (6,9) | 10 (13,5) | <0,023* |
|  | No | 19 (14,3) | 22 (16,2) | 18 (16,8) | 17 (17,9) | 19 (26,4) | 24 (32,4) |  |
|  | I don't know | 91 (68,4) | 89 (65,4) | 73 (68,2) | 69 (72,6) | 48 (66,7) | 40 (54,1) |  |

According to the results, 33% of the participants were familiar with the SUDEP term. These respondents (n=384) further answered the question about through which source they had heard about SUDEP. The sources of awareness of the participants were physicians (27.6%), the internet (20.3%), written and visual media (8.3%), and other sources (43.8%) (Figure 2). 86.9% of the participants stated that they did not have knowledge about SUDEP. Only 2.6% of the remaining students said that they had a knowledge of SUDEP. In addition, 82% of medical students mentioned that they wanted to broaden their knowledge about SUDEP (Figure 3).

**Fig. 2.**
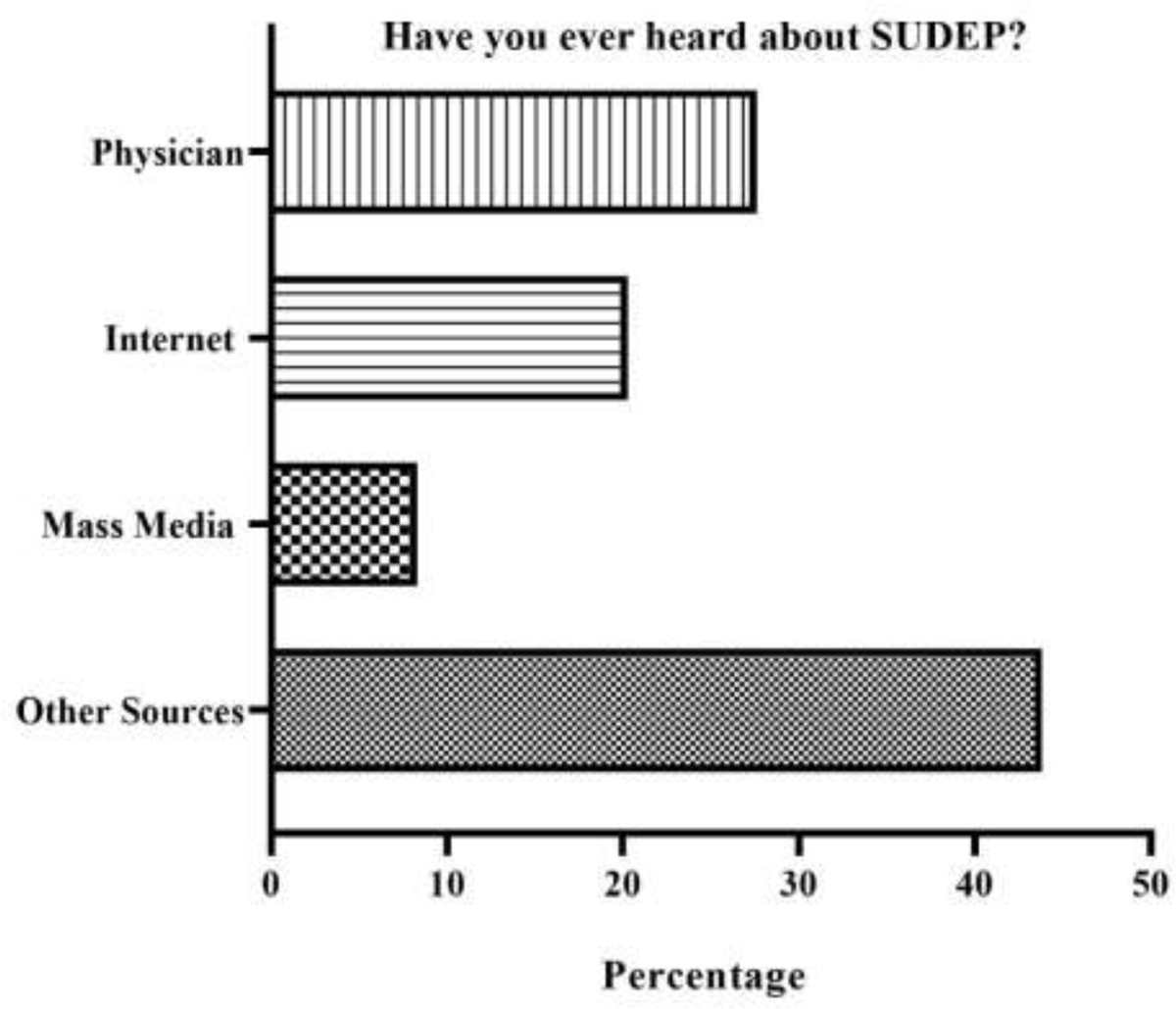
Knowledge of SUDEP. The minority of medical students (33%) have already heard about SUDEP. Source of their knowledge comes mostly from physicians and other sources.

**Fig. 3.**
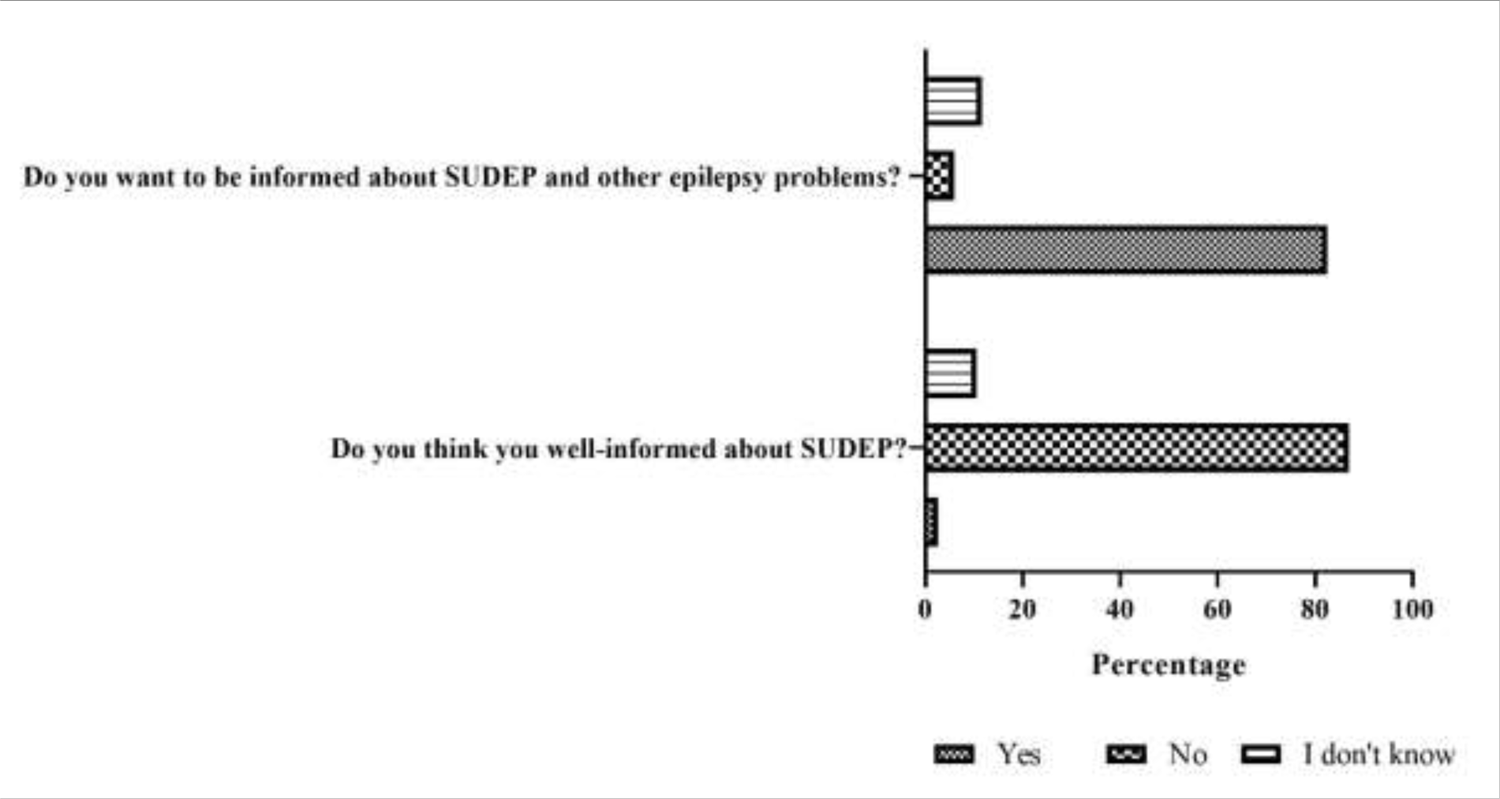
Medical students’ own awareness and willingness to learn more about SUDEP. Although the majority of the participants evaluate themselves to be not well informed about SUDEP, they have interest in learning more about SUDEP and epilepsy.

In order to determine the ability of medical students to define and evaluate SUDEP, the survey included a question about the effect of epilepsy disease on quality of life. 49.4% (n=394) of the students evaluated the effect of epilepsy on quality of life as high, 28.9% (n=229) as moderate, and 17.9% (n=142) as very high (Figure 4, Table 4). In the questionnaire, medical students were also asked about the correct sleeping position for epilepsy patients which is crucial factor for quality of life. 41.6% (n=331) of the participants answered correctly by choosing the side position (Figure 5).

**Fig. 4.**
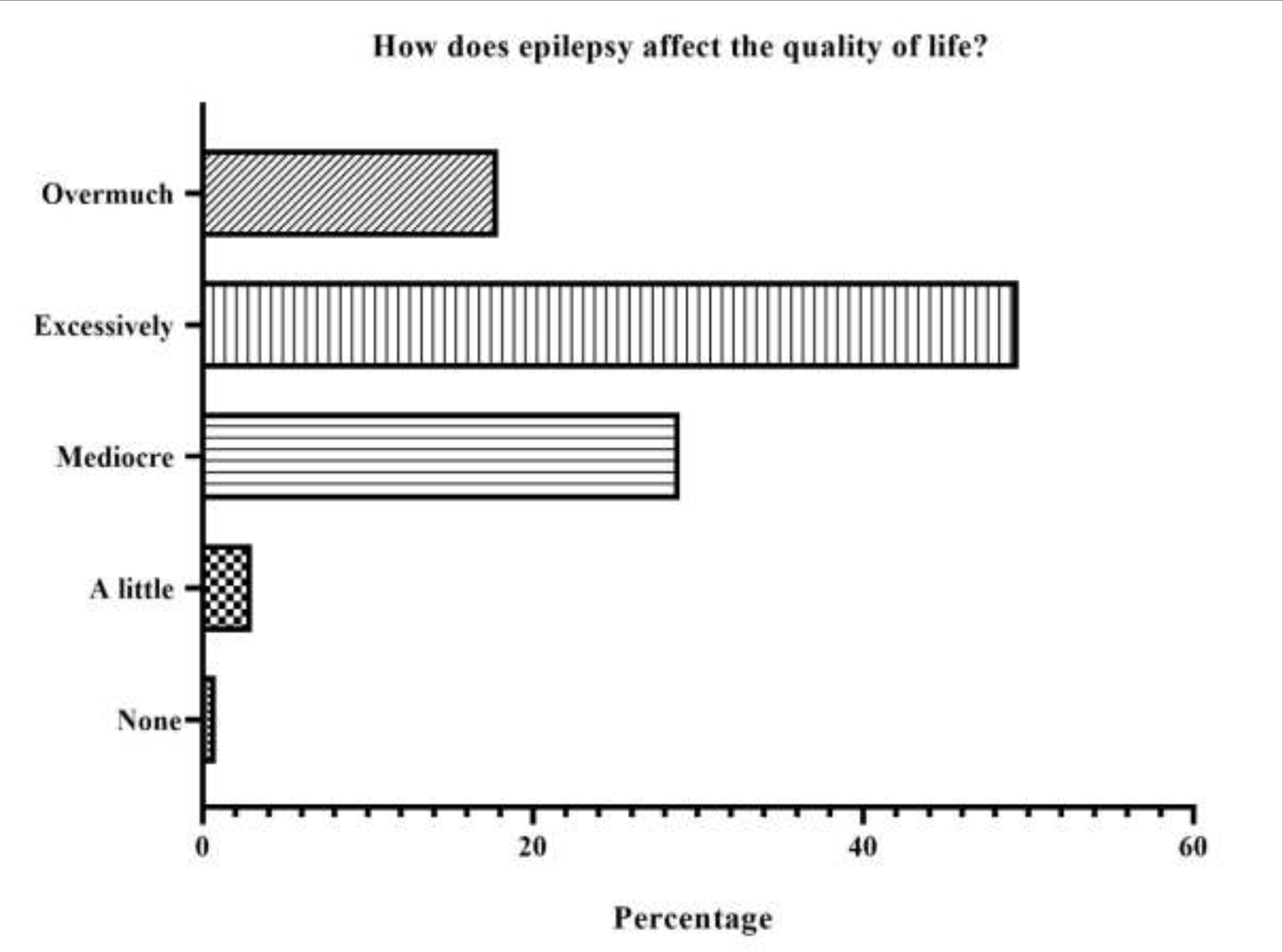
Effect of epilepsy on quality of life based on the medical students’ answers. Most of the medical students consider epilepsy having excessive effect on quality of life.

**Fig. 5.**
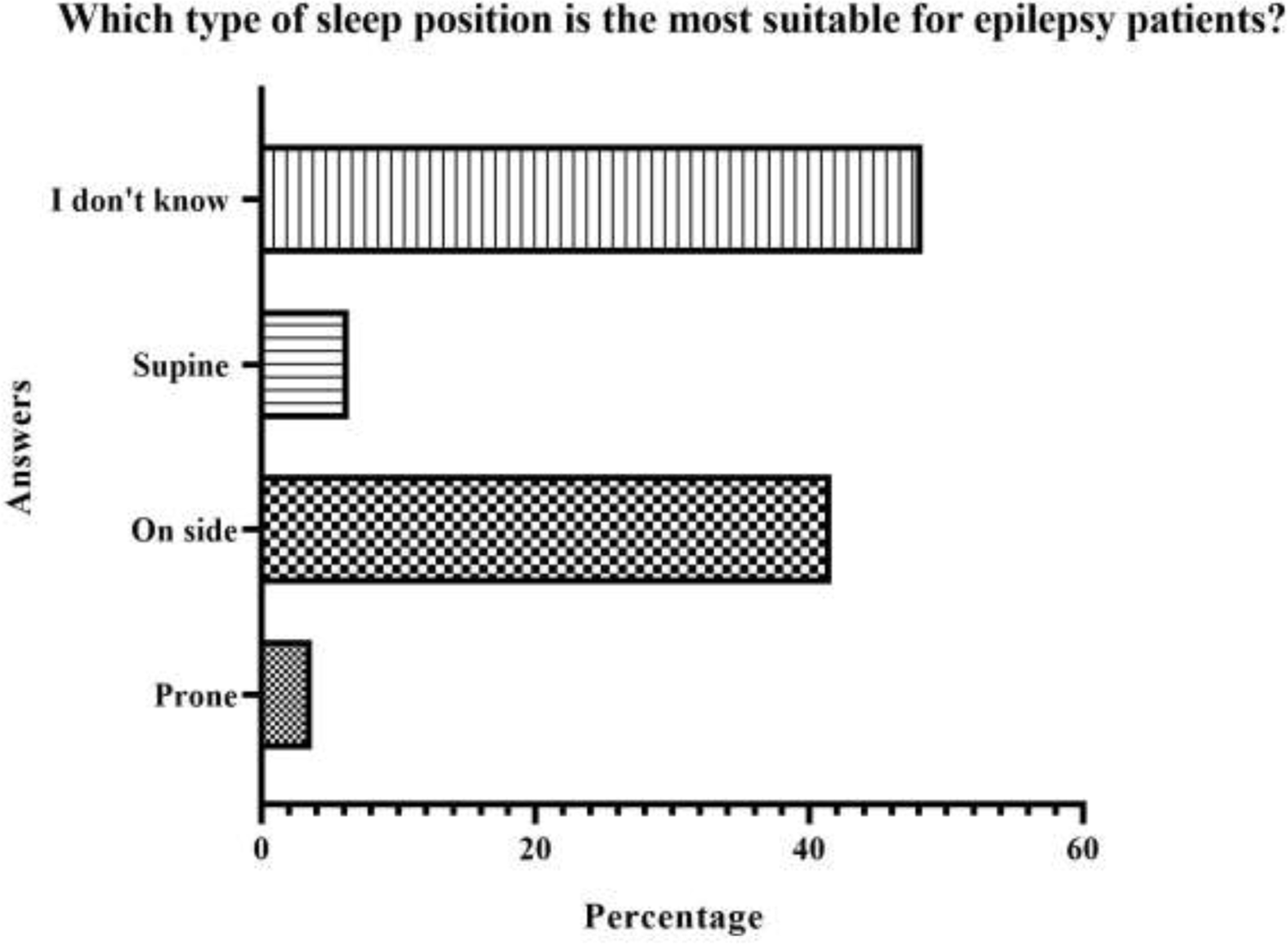
Proper sleep position for epilepsy patients based on respondents’ answers. Although most of the medical students have no information about correct sleeping position for epilepsy patients, nearly half of them chose the side sleeping position which is recommended for epilepsy patients.

**Table 4.** Knowledge of Epilepsy and SUDEP regarding questionnaire according to the term.

| Questions | Answers | Grade 1(n/%) | Grade 2(n/%) | Grade 3(n/%) | Grade 4(n/%) | Grade 5(n/%) | Grade 6(n/%) | P |
| --- | --- | --- | --- | --- | --- | --- | --- | --- |
| **How much does epilepsy affect life?** | None | 2 (1,2) | 0 (0) | 4 (2,7) | 0 (0) | 0 (0) | 0 (0) | <0,149 |
|  | A little | 4 (2,4) | 7 (4) | 6 (4,1) | 2 (1,5) | 1 (1,1) | 4 (4,5) |  |
|  | Mediocre | 47 (28,7) | 43 (24,9) | 38 (26) | 43 (33,1) | 27 (29,3) | 31 (35,2) |  |
|  | Excessively | 82 (50) | 81 (46,8) | 73 (50) | 66 (50,8) | 51 (55,4) | 39 (44,3) |  |
|  | Overmuch | 29 (17,7) | 42 (24,3) | 25 (17,1) | 19 (14,6) | 13 (14,1) | 14 (15,9) |  |
| **Have you heard of SUDEP before?** | Yes | 52 (31,7) | 53 (30,6) | 46 (31,5) | 41 (31,5) | 40 (43,5) | 40 (43,5) | <0,355 |
|  | No | 112 (68,3) | 120 (69,4) | 100 (68,5) | 89 (68,5) | 52 (56,5) | 52 (56,5) |  |
| **Do you think you know enough about SUDEP?** | Yes | 2 (1,2) | 6 (3,5) | 3 (2,1) | 3 (2,3) | 5 (5,4) | 2 (2,3) | <0,404 |
|  | No | 148 (90,2) | 146 (84,4) | 126 (86,3) | 115 (88,5) | 79 (85,9) | 75 (85,2) |  |
|  | I don't know | 14 (8,5) | 21 (12,1) | 17 (11,6) | 12 (9,2) | 8 (8,7) | 11 (12,5) |  |

In addition, students were asked about possible ways to prevent SUDEP. Among the students who answered the question, 33 recommended drug therapy, 31 advocated for increasing awareness of SUDEP, and 11 suggested treatments with wearable technologies and seizure control.

## 4. DISCUSSION

In this survey study, we determined the awareness and SUDEP-knowledge of medical students in Turkey. Results revealed that only 33% of the medical students participating in the survey had heard of SUDEP. A study in an epilepsy center in Germany among epilepsy patients demonstrated that more than 87% of the participants had never heard of SUDEP before and there were only four patients with SUDEP information [10]. The clinicians need to inform the patients about SUDEP and risk factors. Accordingly, informing medical students about SUDEP in the early stages of medical education is expected to increase the awareness of epilepsy patients in the future. A study investigating the importance of SUDEP counseling showed that neurologists with long clinical experience provide SUDEP counseling in patients with refractory epilepsy [11].

In this survey, the source of epilepsy knowledge among medical students was also examined. Results showed that the majority of the participants obtained knowledge from different sources including family relatives and acquaintances, as well as the internet and mass media [12]. A previous study demonstrated that the sources for SUDEP knowledge among adult epilepsy patients were epilepsy agencies, neurologists, and the internet [4]. This evidence suggested that epilepsy and SUDEP awareness is derived not only from physicians or health professionals, but also from the internet or similar sources. Our survey showed that 44% of the medical students obtained SUDEP information from other sources while only 28% got it from the physicians. This can be explained that clinical-term students are more frequently exposed to meetings with neurologists in the hospital.

Another study investigated the impact of patients’ knowledge about SUDEP and epilepsy, and it was demonstrated that improving patients’ knowledge and awareness of SUDEP might be beneficial in both sociodemographic and clinical terms [13]. These findings reveal the important value of health education, which is a crucial part of epilepsy management. In our study, 85% of the medical students were unfamiliar with SUDEP. Clinicians should be well-informed about SUDEP even from the early stages of the undergraduate period, in order to be able to effectively and correctly inform patients in the future during their clinical visits.

Studies have also highlighted the importance of the knowledge and awareness of seizures among healthcare professionals. In this regard, a study among healthcare staff in a hospital in China demonstrated that those who knew moderately about epilepsy mentioned that their approach towards patients changed positively after being informed about epilepsy [14]. This evidence suggests that further research is needed to identify the practical aspects and clinical impact of epilepsy knowledge, especially for epilepsy management. In our study, we also investigated the level of awareness of seizures among medical students. Our data showed that unlike the awareness of SUDEP, seizures were known by the majority (90%) of the medical students.

It has been indicated that sleeping in the prone position may not prevent SUDEP in the cases of non-fatal recurrent seizures [15]. However, evading the prone position may reduce the risk of SUDEP in patients who are immobile in the postictal phase. Our survey’s question about the sleep position concluded that 48.3% of medical students had no information about the appropriate sleeping position for patients with epilepsy. As lying on the side during seizures is a recommended approach, 41.6% of the students thought that epilepsy patients should lie on their side, regarding the ratio of students who are uninformed. In addition to this, neurologists rarely discuss SUDEP with their patients/caregivers. Negative reactions and underestimation of the risk of SUDEP may result in suggestive lack of awareness among practitioners, patients, and caregivers [16]. Framing the discussion in a positive way and using different resources to support patients/caregivers around SUDEP can minimize negative reactions.

Increasing the studies to raise awareness about the sleeping position is essential. Studies have shown that psychosocial factors including emotion regulation disorder and stigma affect the quality of life of patients with epilepsy [17]. Therefore, psychological interventions can be recommended as additional treatment strategies to improve the quality of life. In our study, almost half of the medical students claimed that epilepsy affects the quality of life severely, highlighting the increased awareness among medical students of the impact of epilepsy in the daily life of epilepsy patients.

After the examining the incidence of cardiac fibrosis in SUDEP cases, cardiac pathology was less severe than sudden arrhythmic death cases, but similar to trauma and epilepsy controls (18). This study showed us the presence and severity of cardiac pathology was greater among SUDEP cases compared to healthy cases. Also, investigation of the effects of age, gender, and sertraline administration on seizure-induced respiratory arrest in a DBA/1 mouse model of SUDEP indicated that cases close to human SUDEP provided evidence of both cardiac and respiratory disorders (19, 20). Accordingly, there is evidence that both events occur in the DBA/1 mouse model of SUDEP (21, 22). Both major respiratory and cardiac abnormalities occur in the SUDEP mouse model may also occur in human SUDEP cases.

Awareness of SUDEP is crucial to prevent patients with epilepsy from facing any possible unexpected situations and to have sufficient knowledge not only of physicians but also of students. The fact that SUDEP is not examined and discussed at the expected level in the literature indicates that the issue should be addressed. Future studies can make changes about SUDEP in the literature, and with this, SUDEP can take its place in the literature.

An important limitation of our study is the fact that our sample may not be a reliable representation of the medical students worldwide. Selection bias, including the internet use may have affected our results. The use of self-reported measures and the potential for recall bias of respondents may also be some other limitations [23]. In addition, the dichotomous nature of the possible answers (yes or no) does not allow for quantitative analysis of the results. Additionally, although the survey can only be filled out once from one account, it can affect the results as it can be filled out several times from different accounts. This may be a limitation of the study by leading to bias.

In conclusion, although the majority of the participants know about epilepsy, their level of knowledge about SUDEP was found to be insufficient. Besides, most of the participants think that they should be educated about SUDEP. These results show lack of awareness of SUDEP in medical education that may pose risks to epilepsy patients and contribute to their relatives’ inability to implement preventive measures. A better understanding of the underlying causes and appropriate educational strategies are needed to overcome this situation. Therefore, future studies are needed to determine the more effective approaches to increase the awareness of SUDEP among medical students. According to our results, preclinical medical students’ awareness of epilepsy is lower than that of preclinical medical students. We assume that the reason for this is that preclinical education curriculum does not cover epilepsy awareness. In general, according to studies, it is assumed that even if medical students hear about epilepsy, the reason for the lack of knowledge and bad attitude about epilepsy is mostly due to the fact that neurological diseases are taught by an internist rather than a neurologist [24].

## Author contributions

All Authors conceived the project, discussed the results, and contributed to the final manuscript.

## Data Availability

All data produced in the present work are contained in the manuscript.

